# Collaborative intelligence in AI: Evaluating the performance of a council of AIs on the USMLE

**DOI:** 10.1101/2025.02.17.25322388

**Authors:** Yahya Shaikh, Zainab Asiya, Muzamila Mushtaq Jeelani, Aamir Javaid, Tauhid Mahmud, Shiv Gaglani, Michael Christopher Gibbons, Minahil Cheema, Amanda Cross, Denisa Livingston, Elahe Nezami, Ronald Dixon, Ashwini Niranjan-Azadi, Saad Zafar, Zishan Siddiqui

## Abstract

The variability in responses generated by Large Language Models (LLMs) like OpenAI’s GPT-4 poses challenges in ensuring consistent accuracy on medical knowledge assessments, such as the United States Medical Licensing Exam (USMLE). This study introduces a novel multi-agent framework—referred to as a "Council of AIs"—to enhance LLM performance through collaborative decision-making. The Council consists of multiple GPT-4 instances that iteratively discuss and reach consensus on answers facilitated by a designated "Facilitator AI." This methodology was applied to 325 USMLE questions across Step 1, Step 2 Clinical Knowledge (CK), and Step 3 exams. The Council achieved consensus responses that were correct 97%, 93%, and 94% of the time for Step 1, Step 2CK, and Step 3, respectively, outperforming single-instance GPT-4 models. In cases where there wasn’t an initial unanimous response, the Council of AI deliberations achieved a consensus that was the correct answer 83% of the time. For questions that required deliberation, the Council corrected over half (53%) of responses that majority vote had gotten incorrect. At the end of deliberation, the Council often corrected majority responses that were initially incorrect: the odds of a majority voting response changing from incorrect to correct were 5 (95% CI: 1.1, 22.8) times higher than the odds of changing from correct to incorrect after discussion. We additionally characterized the semantic entropy of the response space for each question and found that deliberations impact entropy of the response space and steadily decrease it, consistently reaching an entropy of zero in all instances. This study showed that in a Council model response variability—often viewed as a limitation—could be leveraged as a strength, enabling adaptive reasoning and collaborative refinement of answers. These findings suggest new paradigms for AI implementation and reveal diversity of responses as a strength in collective decision-making even in medical question scenarios where there is a single correct response.

**Author Summary:** In our study, we explored how collaboration among multiple artificial intelligence (AI) systems could improve accuracy on medical licensing exams. While individual AI models like GPT-4 often produce varying answers to the same question—a challenge known as "response variability"—we designed a "Council of AIs" to turn this variability into a strength. The Council consists of several AI models working together, discussing their answers through an iterative process until they reach consensus.

When tested on 325 medical exam questions, the Council achieved 97%, 93%, and 94% accuracy on the Step 1, Step 2CK, and Step 3, respectively. This improvement was most notable when answers required debate: in cases where initial responses disagreed, the collaborative process corrected errors 83% of the time. Our findings suggest that collective decision-making— even among AIs—can enhance accuracy and AI collaboration can potentially lead to more trustworthy tools for healthcare, where accuracy is critical. By demonstrating that diverse AI perspectives can refine answers, we challenge the notion that consistency alone defines a "good" AI. Instead, embracing variability through teamwork might unlock new possibilities for AI in medicine and beyond. This approach could inspire future systems where AIs and humans collaborate (e.g. on Councils with both humans and AIs), combining strengths to solve complex problems. While technical challenges remain, our work highlights a promising path toward more robust, adaptable AI solutions.

## Introduction

Since the release of OpenAI’s Generative Pretrained Transformer 3.5 (GPT 3.5) in December 2022, many studies have evaluated the performance of Large Language Models (LLMs) on medical knowledge and licensing exams,(1–21) While performance has improved across GPT model updates, varying performance has been noted when the same question is asked to a LLM multiple times. (22,23) This is due to the probabilistic token-by-token generation of LLM content, which can generate a variety of responses to the same question, some of which are incorrect or ‘hallucinations’.(24) Response variability represents the presence of multiple linguistic, though not necessarily factual, reasoning paths for a given question. This suggests the possibility that intersecting multiple reasoning paths may result in re-shaping of each other’s reasoning. And it raises the question of how well an artificial intelligence collective of intersecting reasoning paths might perform on medical knowledge and licensing exams.

In this study, we developed a method to create a Council of AI agents (a multi-agent Council, or ensemble of AI models) using instances of OpenAI’s Generative Pretrained Transformer 4 (GPT4) and evaluate the Council’s performance on the United States Medical Licensing Exams (USMLE).

## Methods

### Sampling the Response Space and Adjudicating Diverse Responses

OpenAI’s GPT-4 was selected as the base LLM given its accessibility, support of Application Programming Interfaces (APIs), extensive documentation and community of support. Our goal was to use a ‘Council’ approach where each LLM instance is a member of the Council, and diverse responses from each LLM instance would undergo a coordinated and iterative exchanges designed to arrive at a consensus response among the models (hitherto referred as “deliberation” or “deliberative process”). To facilitate a deliberative process when there are divergent responses, a Facilitator algorithm (which includes instantiations of the LLM) summarizes the reasoning in each response, formulates a question to help the Council deliberate divergent reasonings, and presents the summary and question to the Council with a request to re-answer the original test question.

The process for the Council’s discussions is summarized below and in Figure 1:

1. The multiple-choice question is copy and pasted into an interface.
2. The question is transmitted to a facilitating algorithm. The facilitating algorithm sends the same question to LLM-communicating functions that represent unique instantiations of the LLM (i.e. unique AI agents), thereby eliciting a response from each LLM instance / AI agent.
3. Each LLM-communicating function communicates uniquely with the LLM through an API.
4. Each instantiation of the LLM sends a response back, generated from its sampling of the response space.
5. Once all the responses are received, they are passed to the Facilitator algorithm, which includes an instantiation of the LLM which assesses Council member response for agreement.
6. If there is no consensus in the responses received, then the Facilitator algorthm does the following: (1) summarizes the responses from the various AI Agents, (2) formulates questions to elicit reasoning relating to the divergent responses, and (3) requests a response to the original question.
7. The output of the Facilitator algorithm is a prompt, which includes a summary of each AI agent’s response and reasoning, a clarifying question relating to differences in response selection, and the initial USMLE question along with multiple choices from which the LLM is required to make a selection.

**Figure 1.**
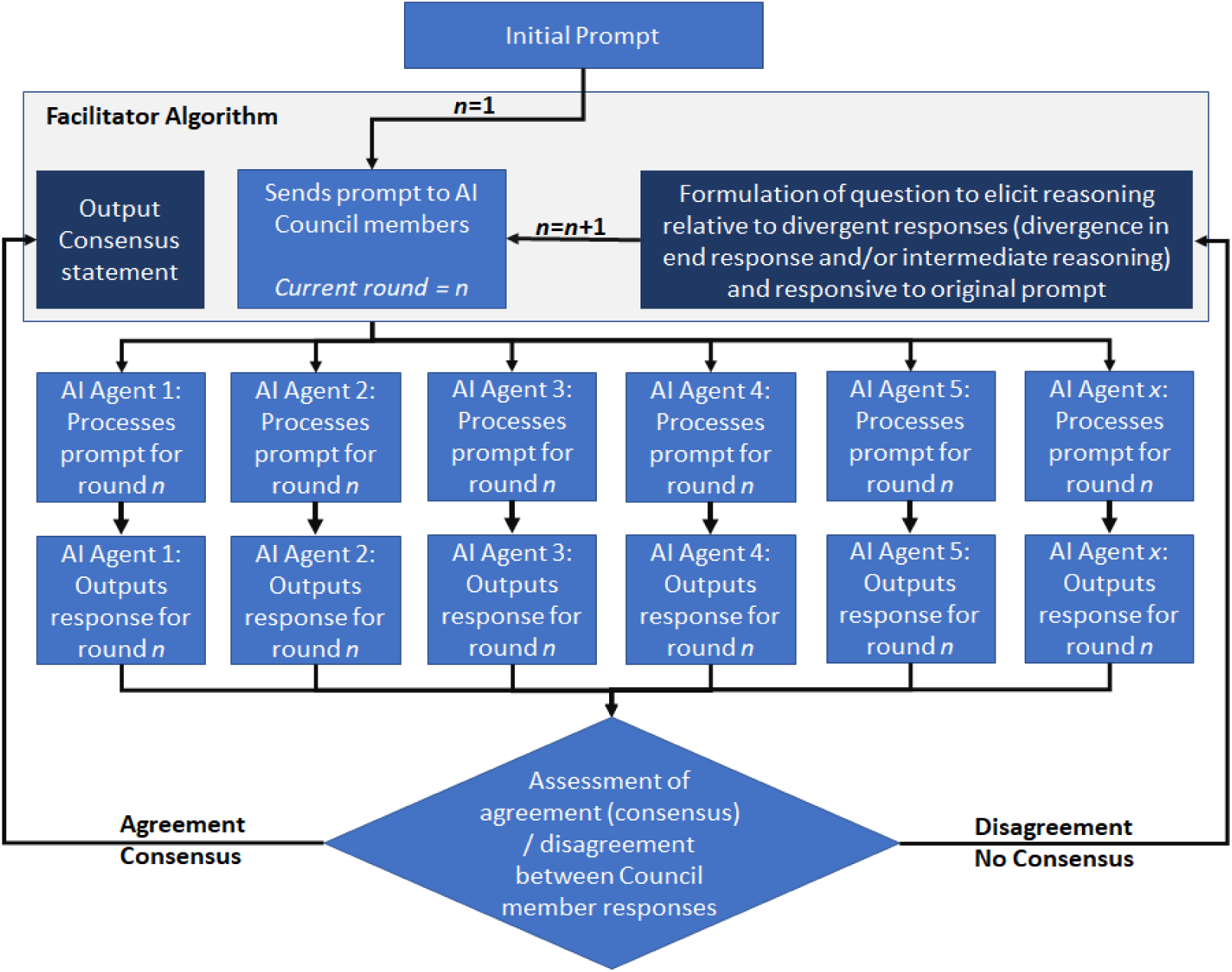
Council of AI Agents Architecture. The User (Human) presents a query, which is passed by the Facilitator algorithm to each of the Council members, whose response is then assessed for agreement. If there is agreement on the answer, a consensus is reached and a consensus statement is output summarizing explanations and identifying the selected response. If there are differing opinions about the correct response, then the Facilitator algorithm summarizes the reasoning presented by each council member, formulates a question that can help clarify reasoning behind difference of opinions, and asks the council members to also respond to the initial prompt (i.e. the USMLE question).

Steps 2-7 are iterated until a consensus response is reached amongst the various instantiations of the LLM. Once a consensus is reached, the transcript of the Council’s deliberations is saved, the server is reset, and the browser is refreshed in order to establish an empty context history for the next question.

For a question requiring deliberation, Table 1 provides a specific example of the Facilitator algorithm’s summarization of Council members’ responses, formulation of question to elicit reasoning related to divergent responses, and a request to respond to the original question. Table 1 does not include the details of the responses from Council members, which can be found as question 25-1 in S2 (S2 Data Raw data files - transcripts of deliberations) as part of transcripts generated by the deliberations conducted by the Council for each question. For details of the API prompts, please refer to the supplemental methods (S1 Supplemental details on methods), which includes prompts for each AI agent, analyzing responses, re-prompting from the Facilitator algorithm to the Council if there is disagreement, and synthesizing responses when there is agreement. All LLMs instances were instantiated with a temperature setting of 1. Code used in this paper is publicly available at: https://github.com/councilofai.

**Table 1.** Example of Facilitator AI’s Question to Council for Discussion in Case of a Divergent Response.

|  |  |
| --- | --- |
| **Original Test Question** | A healthy 30-year-old woman participates in a study of the relationship between cardiovascular function and posture. Placed in a room with a comfortable temperature (72°F) and humidity (40%), she moves from a standing to a supine position. Which of the following sets of changes is most likely in the reflex control of this woman's cardiovascular system?<br>(A) Carotid Sinus Baroreceptor Activity, Atrial Stretch Receptor Activity, and Vagal Efferent Activity to the Heart increase (B) Carotid Sinus Baroreceptor Activity and Atrial Stretch Receptor Activity increase, while Vagal Efferent Activity to the Heart decrease (C) Carotid |
|  | Sinus Baroreceptor Activity and Vagal Efferent Activity to the Heart increase, while Atrial Stretch Receptor Activity decrease (D) Carotid Sinus Baroreceptor Activity increase, while Atrial Stretch Receptor Activity and Vagal Efferent Activity to the Heart decrease (E) Carotid Sinus Baroreceptor Activity decrease, while Atrial Stretch Receptor Activity and Vagal Efferent Activity to the Heart increase (F) Carotid Sinus Baroreceptor Activity and Vagal Efferent Activity to the Heart decrease, while Atrial Stretch Receptor Activity increase (G) Carotid Sinus Baroreceptor Activity and Atrial Stretch Receptor Activity decrease, while Vagal Efferent Activity to the Heart increase (H) Carotid Sinus Baroreceptor Activity, Atrial Stretch Receptor Activity, and Vagal Efferent Activity to the Heart decrease |
| **Facilitator's Prompt to Council for Discussion of Divergent Responses** | AI_A, AI_B, and AI_C all agree that transitioning from a standing to a supine position would result in an increase in carotid sinus baroreceptor activity, atrial stretch receptor activity, and vagal efferent activity to the heart. They argue that the increase in blood volume and pressure due to the change in position would stimulate the carotid sinus baroreceptors and atrial stretch receptors. This would then trigger an increase in vagal efferent activity to slow the heart rate and maintain blood pressure.<br>AI_D, however, suggests that while atrial stretch receptor activity and vagal efferent activity would increase, carotid sinus baroreceptor activity could potentially remain stable or decrease if blood pressure is successfully maintained by these changes. AI_E proposes a similar |
|  | argument to AI_D, suggesting that carotid sinus baroreceptor activity would decrease due to less demand for compensation in blood pressure, while atrial stretch receptor activity and vagal efferent activity would increase. Key Differences: The key difference in perspectives lies in the expected change in carotid sinus baroreceptor activity. AI_A, AI_B, and AI_C predict an increase due to the rise in blood pressure from the positional change. In contrast, AI_D and AI_E predict a decrease or stability in carotid sinus baroreceptor activity, arguing that the body's successful maintenance of blood pressure would reduce the need for these baroreceptors to compensate. Question to Clarify Disagreements: Can the body's maintenance of blood pressure during a transition from a standing to a supine position potentially reduce the need for carotid sinus baroreceptor activity, or would the increase in blood pressure inherently stimulate these baroreceptors regardless of the body's compensatory mechanisms? Based on your preceding response, how would you answer the original question? |

### Selecting the Number of Council Members

There are several technical constraints that determine the number of instantiated AI agents. In the implemented architecture, the LLM’s synthesis of responses from the members of the Council means that all of their responses are passed together within a single API call. The size of the content passed in a single API call is constrained by token limits of a given LLM. Another constraint is the time taken to respond to each call increases linearly by the number of API calls that are made. The latency time between call and response may be quite large for models like GPT-4. Cost is an additional constraint that increases linearly with the number of instantiated AI entities. We found that generating five AI agents as part of the Council allowed us to stay within token limits during Council deliberations.

### USMLE Questions

The USMLE questions used in this study have previously been used in single-AI evaluations of OpenAI’s GPT LLM,(2,4) initially sourced from the June 2022 release of the sample exam by the Federation of State Medical Boards and the National Board of Medical Examiners.(25) The ChatGPT-4 model used in this study was released in March 2023 and was trained on web data available up until September 2021.(26) Because the training cut-off for GPT-4 version used (September, 2021) was earlier than the release of the sample exam questions (June, 2022), it ensured that the multiple-choice questions used to assess the Council of AI were not previously seen in the training data of the underlying LLM. From the original 376 questions that were made publicly available, 51 questions containing images or tables were removed from the questions bank, leaving 325 multiple choice questions (Step 1: 94, Step 2CK: 109, Step 3: 122).

### Analysis of AI Council Performance

We assessed the accuracy of the individual Council members’ initial responses and the Council’s final consensus responses. We also evaluated the relationship between the number of incorrect initial responses and the accuracy of the consensus responses, and the mean number of rounds needed to reach a correct consensus. We calculated semantic entropy of the response space, which measures the uncertainty in a model’s output by evaluating the diversity of meanings among its possible responses.(27) A higher semantic entropy indicates greater uncertainty, suggesting a wider range of potential meanings in the model’s response space. Because we did not have direct access to a model’s output probabilities, we approximated semantic entropy by calculating discrete semantic entropy applying the Shannon entropy formula to the distribution of cluster probabilities:(27)

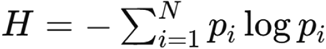

where 𝑁 is the total number of clusters, and 𝑝_𝑖_ is the probability of the 𝑖-th cluster.

We calculated semantic entropy at the end of each round to assess if and how it changes over rounds, and if there is a relationship between number of rounds to reach consensus (semantic entropy of 0) and magnitude of semantic entropy at the end of the initial round.

Because a consensus in round 1 of the Council’s responses indicate an unanimous majority response, we further analyzed the effectiveness of group discussion compared to majority voting for questions where the majority response was not initially unanimous. For each question that was not initially unanimous, we conducted a contingency analysis for the correctness of the majority vote at the end of round 1 versus the post-discussion consensus. For these paired observations, we calculated the matched pairs odds ratio and utilized the McNemar’s test with continuity correction to assess the significance of changes in correctness before and after discussion.

## Results

The Council’s consensus response was correct 97%, 93%, and 94% of the time on Step 1, Step 2-CK and Step 3, respectively. In examining initial responses to questions (i.e. before any rounds of discussion), all council members provided an initial response that was correct on 79%, 78%, and 77%, respectively. (Table 2) 22% of all questions asked (21% of Step 1, 22% of Step 2CK, 23% of Step 3) required discussion because at least one instance of the LLM suggested an incorrect response. (Table 2)

**Table 2:** Council of AI Performance Overview.

|  | Step 1 |  | Step 2 |  | Step 3 |  | All Questions |  |
| --- | --- | --- | --- | --- | --- | --- | --- | --- |
|  | N | % | N | % | N | % | N | % |
| Total Number of Questions | 94 |  | 109 |  | 122 |  | 325 |  |
| **Final Consensus Response Accuracy** |  |  |  |  |  |  |  |  |
| Correct | 91 | 96.8 | 101 | 92.7 | 116 | 94.3 | 308 | 94.7 |
| Incorrect | 3 | 3.2 | 8 | 7.3 | 6 | 5.7 | 17 | 5.2 |
| **Members' Initial Response Accuracy** |  |  |  |  |  |  |  |  |
| All Council members answer correctly | 74 | 78.7 | 85 | 78.0 | 94 | 77.0 | 254 | 78.2 |
| Majority Vote Correct | 88 | 93.6 | 97 | 90.0 | 113 | 92.6 | 298 | 91.7 |
| At least 1 Council member answered incorrectly (initial round) | 20 | 21.3 | 24 | 22.0 | 28 | 23.0 | 72 | 21.8 |
| All Council members answer incorrectly (initial round) | 1 | 5.0 | 4 | 16.7 | 5 | 17.9 | 10 | 13.9 |
| All Council members unanimously agreed on incorrect response (initial round) | 1 | 5.0 | 3 | 12.5 | 3 | 10.7 | 7 | 9.7 |

Compared to a simple majority voting approach (i.e. a correct response given by majority of Council members in the first round), the Council performed better overall (95% for Council vs 91% for majority vote) and for each Step (97% vs 93%; 94% vs 90%; 94% vs 93% for Council vs simple majority for Step 1, 2-CK, 3 respectively) (Table 2). Transcripts of Council deliberations for each question are provided within the supplemental materials. In a contingency analysis comparing the accuracy of the initial majority vote in Round 1 with the Council’s final consensus (Table 3), the Council rarely reverted from a correct initial majority to an incorrect consensus (occurring in only one instance). The odds of a majority voting response changing from incorrect to correct were 5 (95% CI: 1.1,22.8) times higher than the odds of changing from correct to incorrect after discussion.

**Table 3:** Accuracy of majority response versus council’s consensus response.

|  |  | Consensus after deliberation |  |  |  |
| --- | --- | --- | --- | --- | --- |
|  |  | Correct | Incorrect | OR (95% CI) | p-value* |
| Majority vote at the end of round 1 | Correct | 44 | 2 | Ref | <0.05 |
|  | Incorrect | 10 | 9 | 5.0 (1.1, 22.8) |  |

\*McNemar's test with continuity correction

Among questions where at least one member suggested an incorrect response, there were frequently other members that suggested a correct response (95%, 83%, and 93% of Step 1, Step 2CK, and Step 3, respectively), resulting in a discussion within the Council. (Table 2) An example of the Facilitator algorithm’s prompt to the Council for deliberation of divergent responses is shown in Table 1. For questions that resulted in a deliberation, the Council reached a consensus that was correct 85% of the time for Step 1, 67% of the time for Step 2, 75% of the time for Step 3, and 75% of the time overall. Regardless of the number of members who proposed an initial response that was incorrect, the final consensus response of the Council could still be correct as long as one member proposed a correct response. (Figure 2) When no member initially proposed a correct response, there were no instances where the Council’s consensus response was correct. This was true even if there was a diversity of incorrect responses leading to a discussion to reach a consensus. (Figure 2)

**Figure 2.**
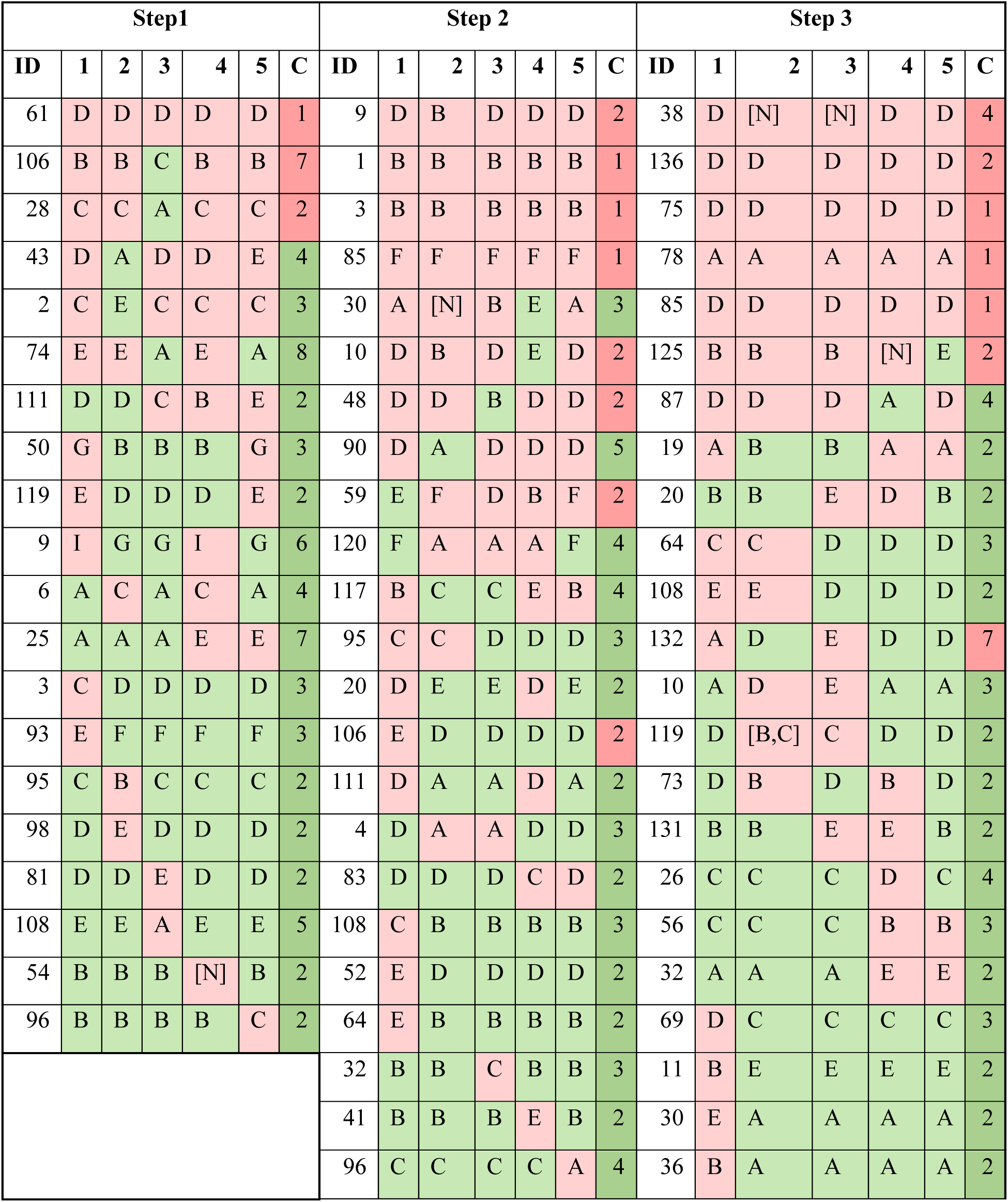
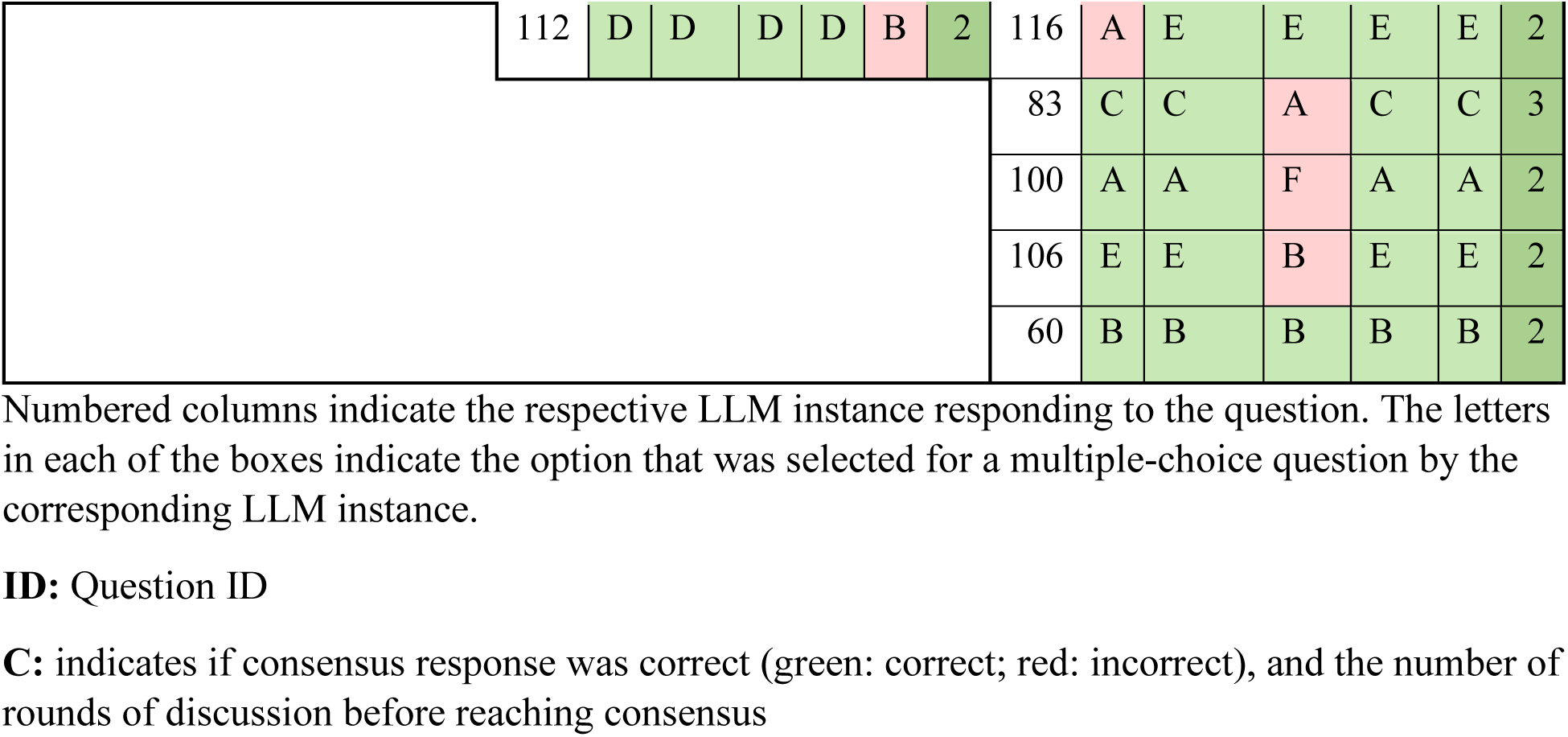
Questions Where at Least LLM Instance Responded Incorrectly in Round 1.

Achieving a consensus response across diverging suggestions required an average of 3.6 rounds of discussion for Step 1, 4.1 rounds of discussion for Step 2CK, and 2.4 rounds of discussion for Step 3. (Figure 3) Of questions requiring discussion, about 85% of Step 1, 71% of Step 2CK, and 72% of Step 3 questions required 2 rounds before reaching consensus. (Figure 4). Of all correctly answered questions, 19% of Step 1 questions, 16% of Step 2 questions, and 18% of Step 3 questions required a discussion before reaching a consensus that was the correct response (Figure 5). There was no significant association (r^2^=0) noted in the number of rounds needed for discussion and the proportion of initial responses that were incorrect. (Figure 6)

**Figure 3:**
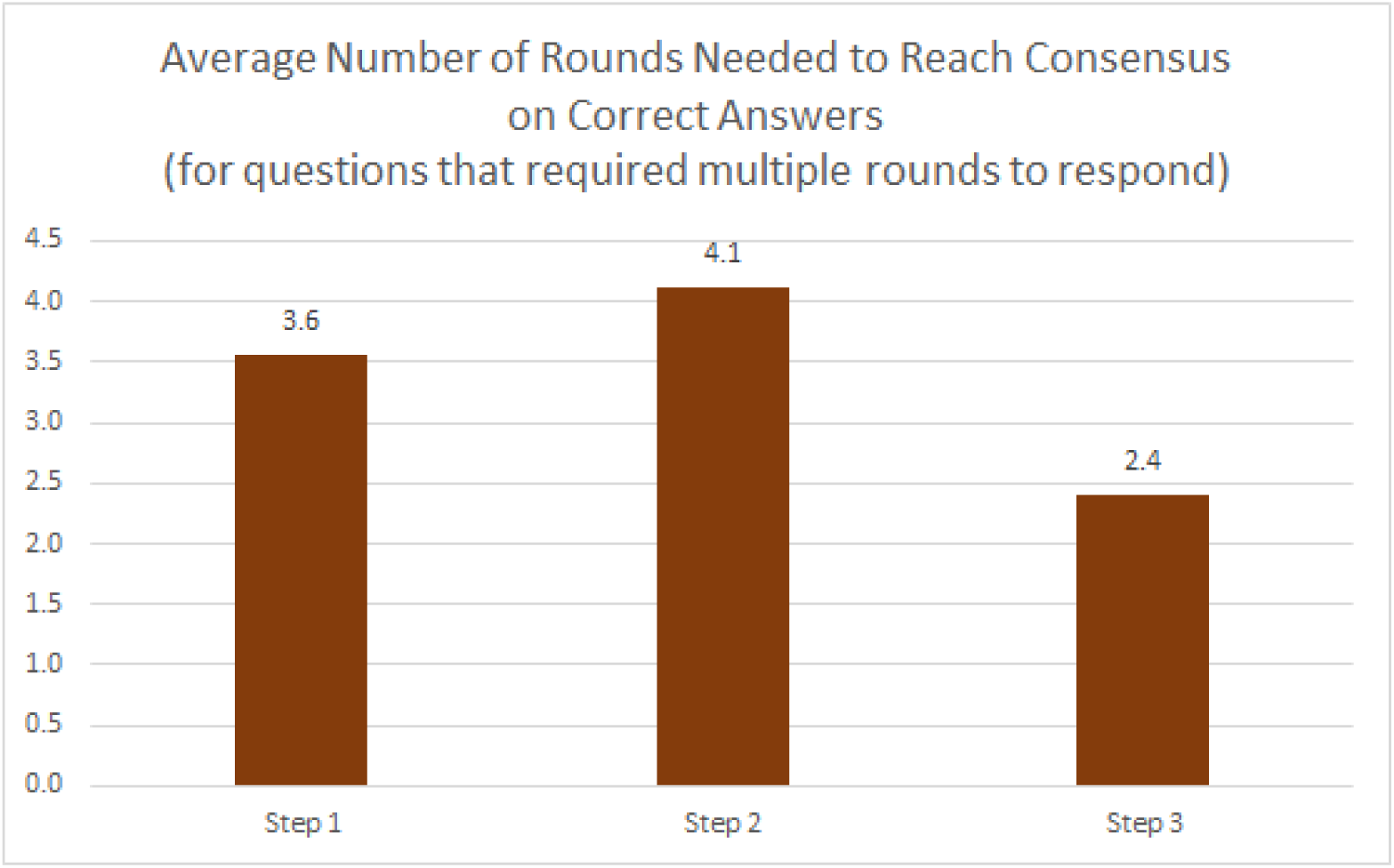
Average number of rounds needed to reach a correct consensus answer.

**Figure 4:**
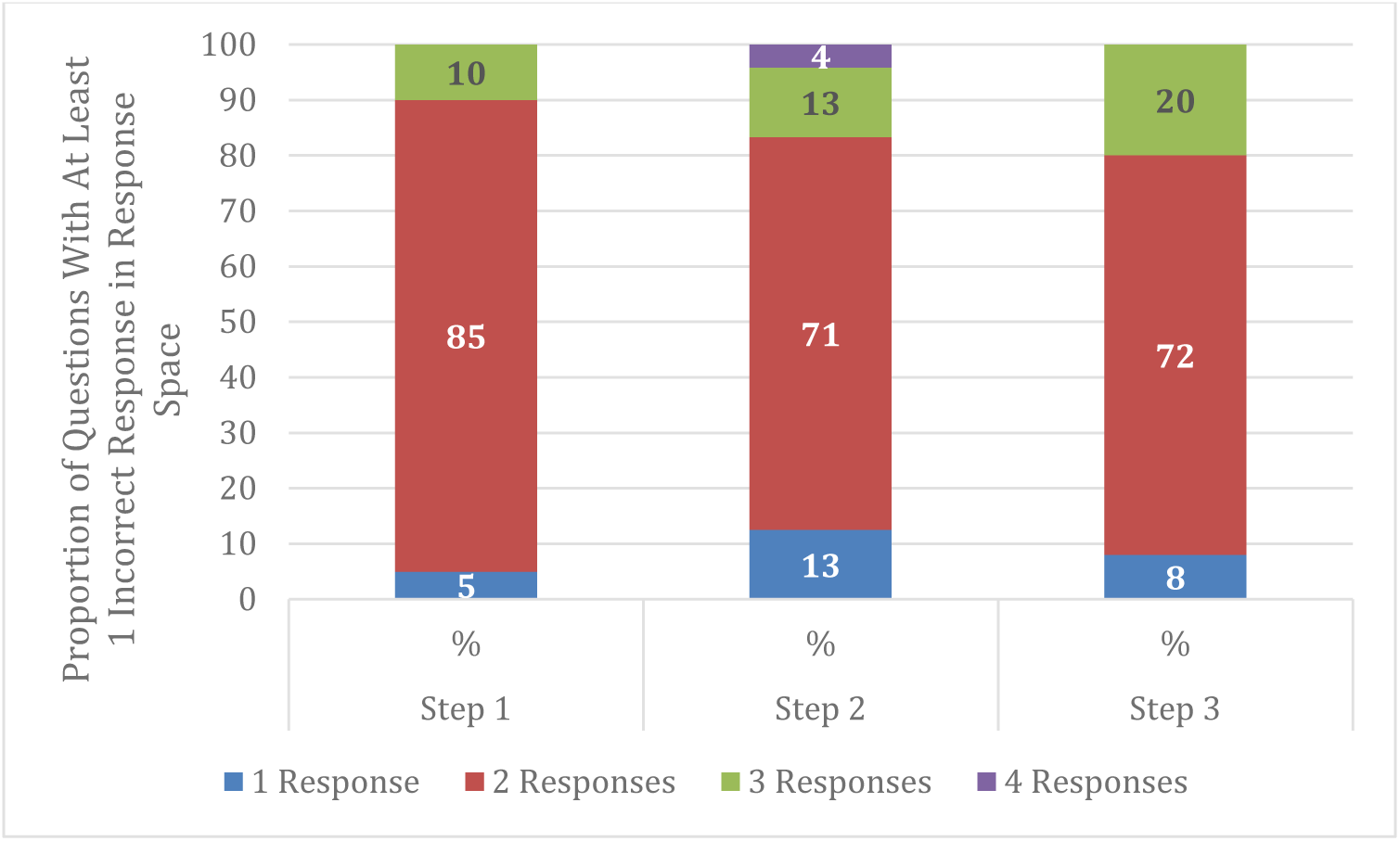
Proportion of Questions with Variability (In Questions With at Least 1 Incorrect Response)

**Figure 5.**
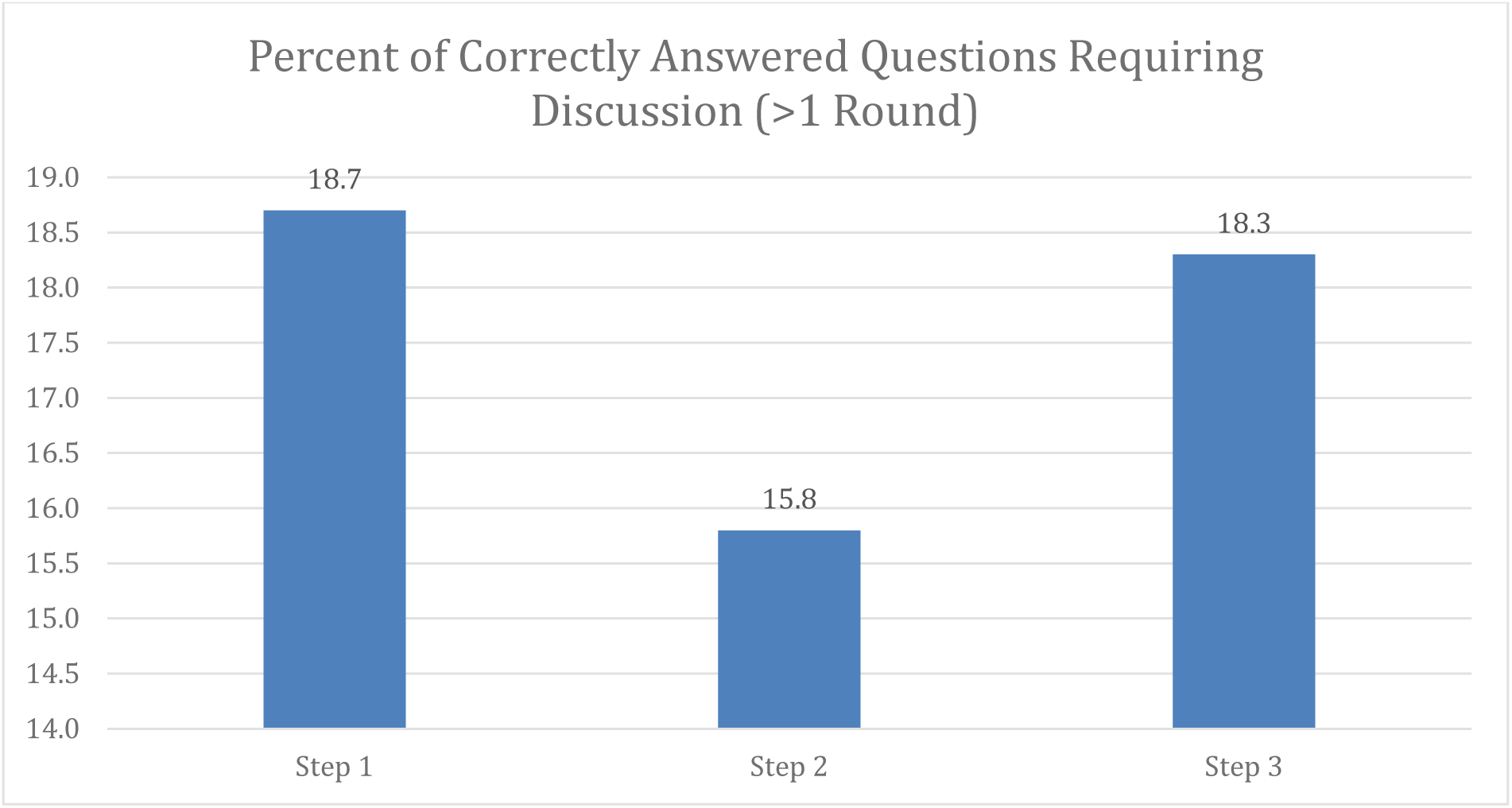
Percent of correct consensus answered questions requiring discussion.

**Figure 6.**
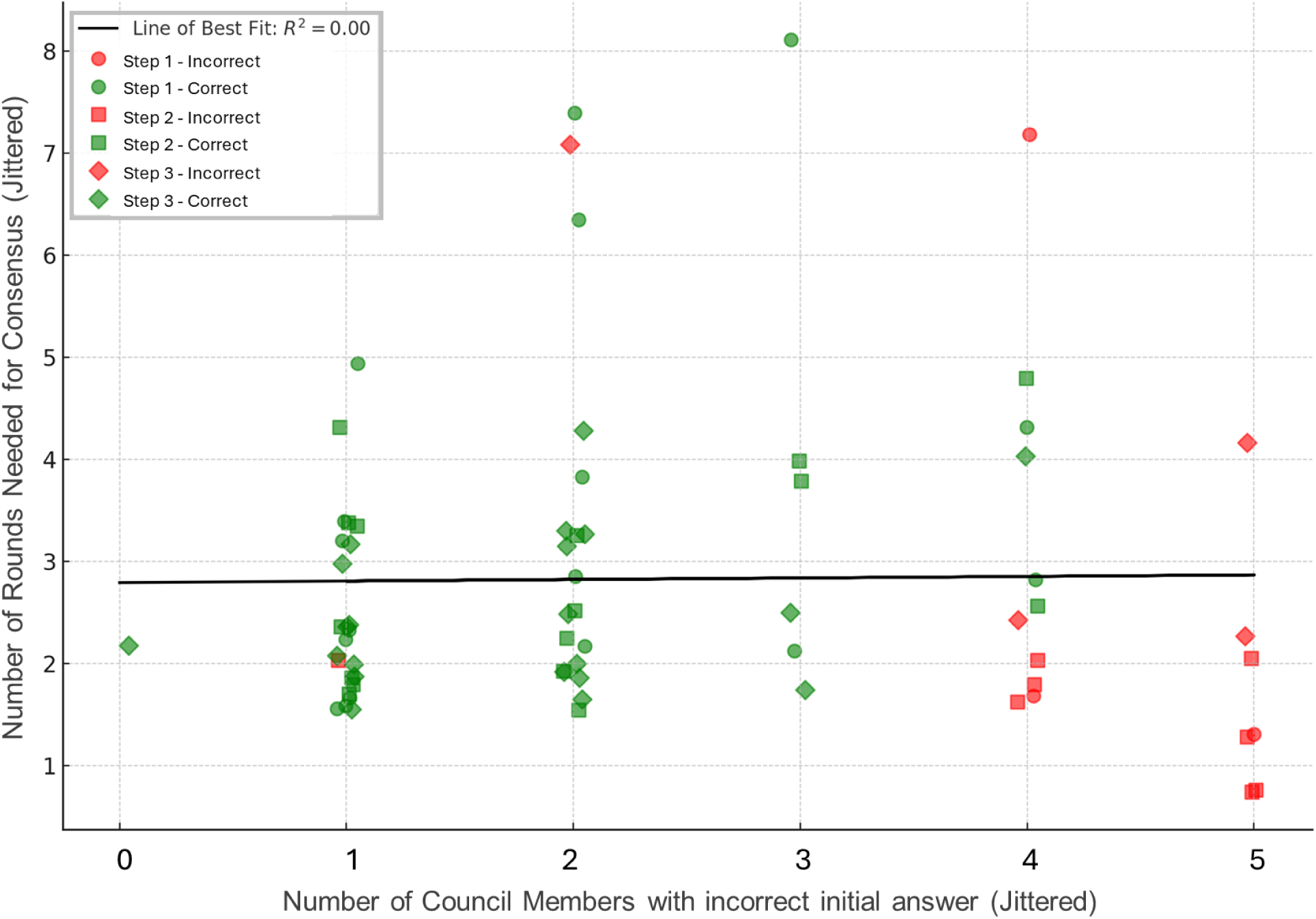
Relationship of number of rounds of deliberation and the number of AI Council Members with an incorrect initial answer. There was no significant association (r2=0) of the number of correct initial answers and the number of rounds of deliberation needed for consensus. A correct consensus answer was only possible when at least one of the initial answers was correct.

We quantified the Council’s degree of internal disagreement at each round of deliberation by calculating semantic entropy, where higher entropy indicates greater divergence in multiple- choice responses and lower entropy indicates growing consensus.(27) Across questions, entropy consistently decreased with each additional re-prompt, reflecting the Council’s steady progression toward unanimity (Figure 7). Regardless of the total number of rounds needed, entropy generally approached zero by the final round. Notably, even in instances where the final consensus was incorrect, entropy still converged toward zero. This suggests that deliberation seems to lead to decreases in entropy of the response space in every case studied, but it does not guarantee accuracy in every case (though it can improve overall accuracy compared to majority vote or single instances of the LLM (Table 2; Table 3)). Furthermore, the number of rounds of deliberation needed to achieve a consensus is not determined by the amount of disagreement in the initial round of responses to a question (Figure 6): entropy of round 1 responses range from 0.2 to 0.3 regardless of the number of rounds needed to reach consensus (Figure 7).

**Figure 7:**
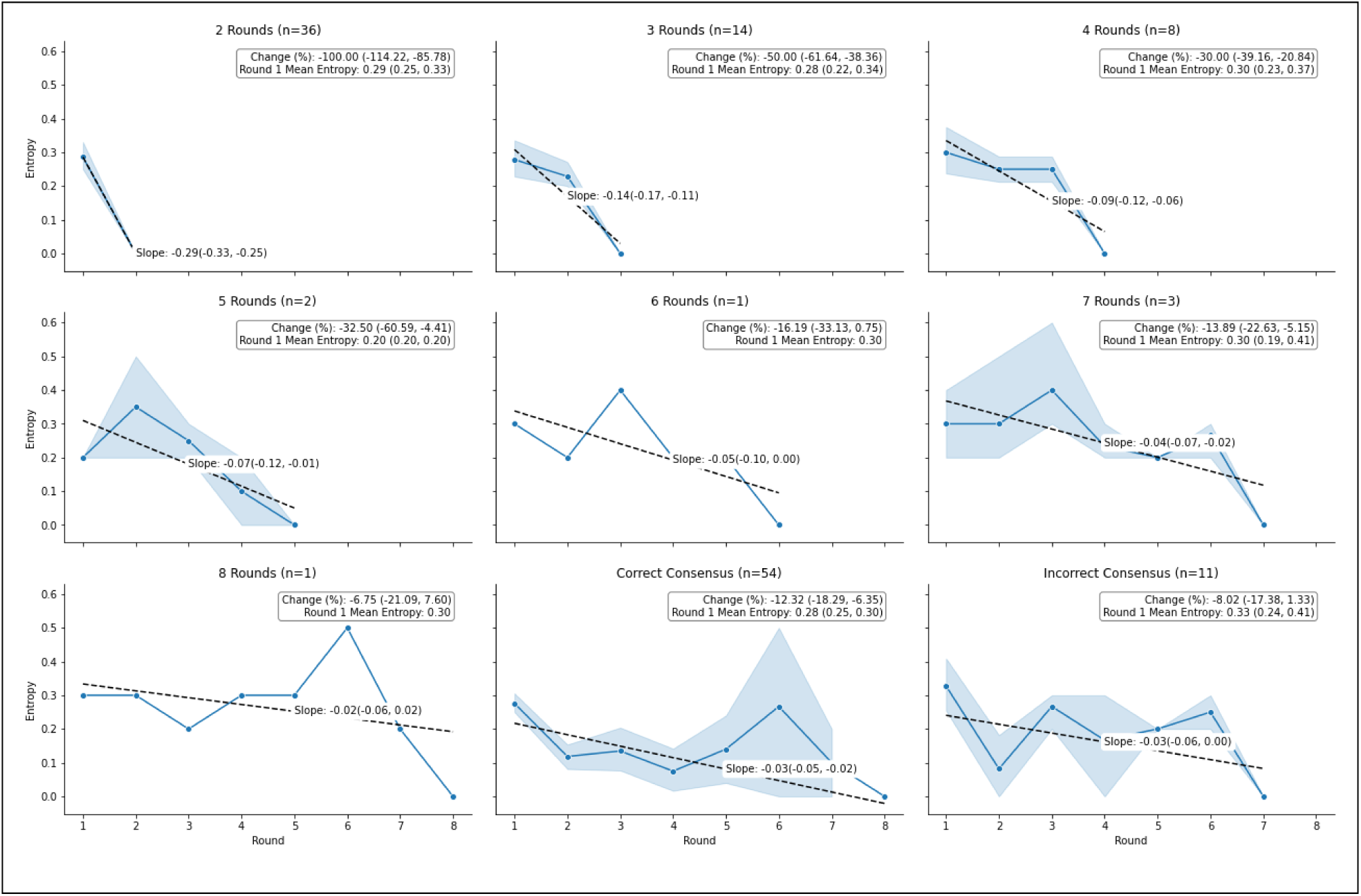
Change in semantic entropy over time by number of rounds of Council deliberation needed for consensus

## Discussion

Collaborative multi-agent approaches are similar to the current practice of medicine, in which input from multiple team members helps to make the best clinical decision for a patient. In this study, we demonstrate the highest ever performance on the USMLE by an AI system through use of a collaborative Council of AI Agents. While a single instance of a LLM (GPT-4 in this case) may potentially provide incorrect answers for at least 20% of questions, a collective process of deliberation within the Council may refine their reasoning pathways,(28) enabling an LLM to correct its incorrect responses 80-90% of the time. This suggests that a multi-agent AI approach can achieve a problem-solving capability on the USMLE unlikely to be achieved by solitary instances and underscores the potential of collaborative AI strategies in medicine.(29) Our review of discussions between AI Council members suggests that collaborative LLMs may better approach the construction of concepts that can be adjusted towards the correct answer, rather than resembling regurgitation from rote memorization as has been previously noted by others.(30) Review of deliberations facilitates human interpretation of linguistic constructions underlying AI decision-making.

### Comparison of USMLE Performance with Prior Studies

Although the exact questions have varied between prior studies of LLMs on USMLE questions, the Council of AI seems to achieve a superior performance to any solitary state-of-the-art LLM to date. In a comparable study utilizing the same USMLE practice questions, GPT-4 answered 88%, 86%, and 90% of Step 1, 2CK, and 3 questions correctly, respectively.(4) In a dataset of USMLE questions known as MedQA, Google’s Med-Gemini-L 1.0 achieved 91% accuracy overall.(31) Prior studies in MedQA have shown an accuracy of 86% by GPT-4 base and 86.5% by MedPaLM 2.(31) By optimizing prompt engineering, Nori et al. were able to increase the performance of GPT-4 to 90% in this dataset.(3) A comparison of the Council of AI to prior zero-shot tests of GPT-4 is shown in Table 4.

**Table 4.** Comparison of Council of AI accuracy on USMLE questions compared to prior studies using GPT-4 (zero-shot)

| Studies using GPT-4 Models (zero shot) | Step 1 | Step 2 | Step 3 |
| --- | --- | --- | --- |
| Council of AI | 97% | 93% | 94% |
| Mihalache et al | 88% | 86% | 90% |
| Nori et al | 81% | 82% | 90% |

AI: Artificial Intelligence. USMLE: United States Medical Licensing Exam.

### The Value of Response Variability

The observation that 22% of questions needed multiple rounds of deliberation highlights the stochastic nature of LLMs’ content generation and suggests that a single response from an LLM might not suffice for precise decision-making. This is consistent with findings from other studies, such as Yaneva et al., who reported variability in correct answers across three replications, observing inconsistencies in 20% of USMLE questions.(32)

This study suggests that variability in LLM responses, a limitation in isolation, can be a strength in collaboration. This unexpected finding suggests new approaches to evaluating generative LLMs, which usually favor predictability and consistency, especially in contexts like professional exams where questions have only one correct answer. By valuing the inherent variability in responses across different LLM instances and leveraging consensus-building as a method, this finding suggests we may need to rethink what optimal behavior means for generative LLMs. Evaluation should not only consider the performance of a single LLM instance but also prioritize the system’s capacity for collaborative engagement with other AI instances to identify a best response. A collaborative approach could significantly improve accuracy and reliability in critical decision-making areas like healthcare.

Interestingly, the Council never achieved a correct consensus response without at least one member offering a correct initial response. This underscores the significance of having a Council with sufficient diversity in reasoning to ensure at least one LLM instance can propose a correct response. Future research could explore the identification of the optimal characteristics, number, and composition of council members to generate a diverse range of responses to increase the probability that at least one member can suggest a correct answer.

An unexpected finding in this study was that the number of rounds of deliberation needed to achieve a consensus was not associated by the extent of disagreement in the initial round of responses to a question (Figure 7). This suggests that a factor other than the number of divergent responses is driving the amount of deliberation. Future studies can explore the characteristics of the question or responses which increase or decrease the amount of deliberation before consensus.

### Self-Correction and Collective Intelligence

If a LLM produced an incorrect initial response, we found it subsequently generated a correct response 85% of the time through interaction with other instances of itself. This aligns with recent studies on self-correction mechanisms in LLMs.(33–35) This phenomenon, where multiple AI instances engage in a dialogue and reconcile their differences to achieve a consensus, reflects a simulation of collective intelligence,(36) enabling a group of AIs to ’change their minds’, a level of complexity and sophistication often attributed to human group dynamics.(37) The necessity for LLM instances to explore new reasoning paths to arrive at a correct consensus underscores the flexibility and adaptive reasoning capabilities present in contemporary LLMs. This flexibility is particularly valuable in dynamic, real-world settings where issues often require more than a singular, predetermined solution strategy.

The results suggest that discussion-based consensus provides a statistically significant improvement in the correctness of responses compared to a simple majority vote. Specifically, in comparing the initial majority vote in Round 1 with the Council’s final consensus (Table 3), there were only two instances where a correct initial majority ultimately resulted in an incorrect consensus after discussion. In contrast, for the 19 questions where the initial majority was wrong, in 10 of those cases the Council consensus overturned the initial majority, resulting in a correct final answer. The odds of a majority vote response changing from incorrect to correct were 5 (95% CI:1.1, 22.8) times higher than the odds of it changing from correct to incorrect after discussion, demonstrating a statistically significant improvement in accuracy from pre-discussion majority voting to post-discussion consensus. Taken together, these findings highlight the Council’s deliberative process as a strategy that preserved correct majority opinions and “rescued” those instances in which the majority initially erred. In other words, the multi-agent Council approach has the potential to both preserve correct majorities and significantly increase accuracy when the majority begins in error. By iteratively reconciling divergent reasoning paths, the Council demonstrates that structured dialogue can leverage variability in AI-generated responses to achieve superior reliability and performance compared to relying solely on a single, one-shot majority vote. Future research might explore *why* discussion can improve accuracy (e.g., type of question, quality of explanations, number of conflicting perspectives), as well as any conditions that optimize its effectiveness.

The high performance of the Council of AI Agents is supported by studies of ensemble systems and multi-agent systems that show that AI collaborations can have a superior performance compared to single AI instances (29,38–48) and can be useful for applications such as drug-discovery.(49,50) Ensemble approaches have also been used to improve performance for LLMs across different knowledge domains. For medical question-answering in particular, Yang et al employs boosting-based weighted majority voting which significantly outperformed individual models on various datasets;(51) Jiang et al uses specific algorithms to merge potential candidate outputs from various LLM in a pair-wise fashion improving performance by measuring against with known benchmarks;(52) Pitis et al introduces a "prompt ensemble" method for a "chain-of-thought" language model reasoning;(53) while Naderi et al uses ensemble techniques for named entity recognition in health and life science domains.(54)

A common communication architecture in a multi-agent system is that of agents that are connected to a middle agent. AI agents that are completing a task may have varying levels of information, with the middle agent (e.g. facilitator, mediator) coordinating between other agents and providing each agent with services that it may need.(39–43) In contrast, an alternative multi-agent architecture, studied by others (55–57) and developed for this study, provides all AI agents with full information, equal guidance for reasoning (i.e. same CoT prompts), and awareness of each other’s responses communicated to them through the Facilitator agent. This architecture simulates a council, where each AI member can respond to a query, and if there is variability in responses, deliberate divergent responses through the Facilitator to reach a consensus response.

Because the deliberation of the Council occurs through a number of different steps, future directions of research can explore the mechanisms of the Council’s response generation, including characterizing and evaluating each stage of the algorithm (i.e. initial response to the USMLE question, the response of the LLM to compare responses for agreement, the response of the LLM to summarization, the response of the LLM to generating a clarifying question, and the subsequent responses of the Council of AI Agents to the new prompt). Additionally, the current study held all LLM attributes (i.e. prompt, temperature, maximum tokens, and top_p) constant between each instantiation to focus the evaluation on the presence of interaction between Council members and the resulting performance. However, a future direction of research can evaluate the impact of modifications to different parameters individually or in concert, with all or part of the Council embodying the changes.

### Limitations

The Council of AI Agents approach, while effective, requires significant computational resources. Future studies could focus on optimizing the efficiency of such systems and exploring their applicability in different domains. Token limits in LLMs are a technical constraint that can limit the depth of analysis, particularly when dealing with complex or lengthy prompts. In our study, we attempted to mitigate this issue through efficient summarization and synthesis of responses by the Facilitator AI, enabling the system to handle a larger number of repeated samples without exceeding token limits.

The exclusive use of OpenAI’s GPT-4 in our Council of AI approach was a strategic choice in this study. Future studies can explore the inclusion of different LLMs, each with unique training datasets, algorithms, and probabilistic models, which may possibly lead to more robust and well-rounded consensus. The variation in the success rate of achieving consensus across different exam steps (Step 1, 2, and 3) suggests challenges in maintaining AI consistency across different contexts and that AI systems might require tailored approaches depending on the specific nature and complexity of the tasks at hand. Testing in other datasets, such as MedQA, may help for more direct comparison to other LLMs in the future. While we selected the present test question set to ensure all questions were from after GPT-4’s training period, a possible limitation is that sample exam questions could be highly similar to older sample exam questions, in which case the LLM may already have encountered highly similar questions in its training data.

Latency time increases with each additional instantiation of the LLM. This presents a significant challenge in real-time decision-making scenarios, especially when multiple rounds of sampling are required. It is possible to eliminate additional latency time while increasing the number of Council members by employing a strategy of synchronous sampling through multiple instantiations of the LLM API, each with different API keys. This approach would allow for parallel processing of responses, substantially reducing the overall time taken to reach a consensus. Through parallel computing, it would be feasible to maintain the depth and quality of analysis without sacrificing response time. For now, this system would work best for non-emergent scenarios that have a tolerance for the time needed by the Council for deliberation and achieving consensus.

Ethical considerations around the use of AI in professional and decision-making contexts need to be addressed, ensuring that the deployment of such technologies is responsible and beneficial to society. A Council of AI approach may facilitate integration of voices of underrepresented communities as council members to try to reduce bias, which may be represented within purposefully trained smaller language models or through fine tuning of LLMs. However, LLMs themselves have an inherent limitation in that they are trained on a written corpus, which excludes communities whose knowledge may be codified in a diversity of non-written media including experiential transmissions of knowledge, oral histories, performative representations, and more. Future studies can explore the extent to which rapidly adopted AI systems are disconnecting communities from their own realities and reshaping community knowledge systems toward more hegemonic representations.(58) Additionally, studies also need to be conducted to formalize AI impact assessment on knowledge systems.

### Conclusions

By designing an algorithm that embraces the variability inherent in LLM responses, the Council of AIs model leverages a multi-agent AI framework to achieve superior performance on the USMLE. This study highlights the importance of collective intelligence and collaborative decision-making in AI systems, offering a model for understanding and maximizing AI capabilities. The Council of AI Agents emphasizes the value of collaboration over individual accuracy and demonstrates the dynamic, evolving nature of AI cognition. This study may provide a framework for one possible future of medical AI, in which multidisciplinary teams of AIs and humans work together to improve health outcomes across the world.

## Data Availability

All relevant data are within the manuscript and its Supporting Information files.

https://github.com/councilofai/project-saru

